# DNA methylation age deviation and cognitive status among older adults in the US, NHANES 1999-2002

**DOI:** 10.1101/2025.09.15.25335796

**Authors:** Erika Walker, Scarlet Cockell, Belinda L. Needham, Andres Cardenas, Sung Kyun Park, Kelly M. Bakulski

## Abstract

Biological aging, measured using DNA methylation, is a potential biomarker for cognitive health outcomes. We evaluated associations between DNA methylation measures of aging and cognition in a nationally representative sample of adults aged 60+ in the National Health and Nutrition Examination Survey (NHANES), 1999-2002. Genome-wide DNA methylation data were used to create 13 measures of biological aging trained on different aging phenotypes. Cognition was assessed with the Digit Symbol Substitution Test (DSST). To evaluate associations between each DNA methylation measure and DSST score, survey-weighted linear regression models adjusted for age, sex, race/ethnicity, education, smoking, serum cotinine, and BMI were run. We assessed effect modification by sex, education, and race and ethnicity. Included participants (N=1,463) were an average of 70.5 years old and 82.7% non-Hispanic White. The average DSST score was 46.9 (SD 17.6). Ten of 13 DNA methylation measures were associated with DSST (adjusted p<0.05). One year of GrimAge2 accelerated aging was associated with –0.41 points lower DSST score (95% CI: –0.61, –0.21; adjusted p=5x10^-4^). In stratified analyses, higher magnitudes of association were observed among male and non-Hispanic White participants across multiple aging measures. DNA methylation may be a useful biomarker of cognitive status among older adults.

## Introduction

Cognitive impairment, reflecting loss of memory, learning difficulties, and difficulty concentrating, impacts approximately 19% of community-dwelling adults over age 50 globally.^1^ Cognitive impairment can reflect specific underlying pathologies that go on to become dementia, or it may reflect age-related cognitive decline among older adults.^2^ To help understand the course of cognition throughout the lifespan, biomarkers are essential. Broadly, biomarkers are measures of biologic characteristics used as indicators of susceptibility/risk, diagnostics, monitoring, prognostics, prediction, pharmacodynamics/response, and safety.^3^ While dementia pathologies such as Alzheimer’s disease have strong diagnostic biomarkers,^4^ earlier indicators of susceptibility or risk, including those associated with poorer cognition among older adults, are more limited.

One promising source for aging related biomarkers is DNA methylation.^5^ DNA methylation variation can reflect underlying genetic sequence, environmental factors, and health status,^6^ which can be integrated into DNA methylation age measures.^7^ Different DNA methylation clocks include chronological, phenotypic, telomere, and mitotic age biomarkers. DNA methylation measures of chronological age, such as by Horvath^8^ and Hannum,^9^ are considered “first generation” clocks that sought to predict chronological age from a biologic specimen. Phenotypic age measures, such as GrimAge,^10^ GrimAge2,^11^ PhenoAge,^12^ and DunedinPoAm,^13^ are considered “second generation” clocks that predict health status or lifespan. Hallmarks of aging including multiple additional metrics^14^ and newer clocks sought to use DNA methylation to estimate telomere length^15^ and cell senescence.^16^ It is common for studies to examine a subset of these DNA methylation aging estimates; however, comprehensive biomarker evaluation requires assessing the utility of DNA methylation age estimates across these different constructs of aging.

DNA methylation aging biomarkers have been associated with cognition,^17,18^ but these studies were often either small or among younger adults,^19,20^ and the types and number of DNA methylation aging biomarkers investigated were more limited. In this study, we sought to build on the existing literature by focusing on older adults and evaluating multiple types of DNA methylation aging biomarkers (chronological, phenotypic, telomere, and mitotic estimates). In a United States nationally representative sample of older adults, we performed a cross-sectional analysis of 13 DNA methylation-based aging biomarkers for association with cognition.

## Methods

### NHANES study population

The National Health and Nutrition Examination Survey (NHANES) is a study operated by the National Center for Health Statistics (NCHS), which is a division of the Centers for Disease Control and Prevention (CDC).^21^ The current format of NHANES, which began in 1999, is an annual, nationally representative, cross-sectional survey of the US civilian population’s health status. Data are collected via interview questionnaires, physical examinations, and biospecimen analysis. The NCHS publicly releases datasets in two-year cycles. This analysis combines data from the 1999-2000 and 2001-2002 cycles (N = 21,004), in which both DNA methylation and cognition measures are available. All NHANES participants provided informed consent. The University of Michigan Institutional Review Board approved this secondary data analysis (HUM00194918). The datasets used in this manuscript are publicly available.

### DNA methylation measures

Participants aged 20 years and older in the 1999-2000 and 2001-2002 cycles were eligible for the original DNA sample collection (N=10,291).^22^ A random sample of participants aged 50 years and older were included in a subset for DNA methylation analysis (N=4,449).^23^ Venous blood was collected and stored at -80**°**C. DNA was extracted and bisulfite converted using the Zymo EZ DNA methylation kit. DNA methylation was measured using the Illumina Infinium HumanMethylation EPIC (version 1) BeadArray on the Illumina iScan. After excluding participants who did not consent to future research use of samples or who had insufficient DNA specimens, the DNA methylation subset included N=2,352 participants.

Full DNA methylation data preprocessing steps are described elsewhere (https://www.n.cdc.gov/nchs/data/nhanes/dnam/NHANES%20DNAm%20Epigenetic%20Biomarkers%20Data%20Documentation.pdf). Briefly, DNA methylation IDAT image files data were processed into methylation intensity values, the proportion methylated at each site (“beta values”) were calculated, probe type bias corrected with BMIQ,^24^ and functional normalization performed.^25^ Samples were excluded with low median intensity on both methylated and unmethylated channels (<10.5 RFU) or with unexpected sex chromosome ploidy. Probes were excluded with detection p-values >1x10^-16^ in at least 1% of samples,^26^ annotated to cross-reactive genomic sites,^27,28^ and annotated to a genetic polymorphism with minor allele frequency >1%.^28,29^

Using the preprocessed DNA methylation data, NHANES calculated and released 13 DNA methylation age measures: Horvath,^8^ Hannum,^9^ SkinBlood,^30^ PhenoAge,^12^ Lin,^31^ Weidner,^32^ VidalBralo,^33^ YangCell,^16^ Zhang,^34^ GrimAge,^10^ GrimAge2,^11^ DunedinPoAm,^13^ and HorvathTelo.^15^ Conceptually, we categorize these into measures of chronological age (Horvath, Hannum, SkinBlood, Lin, Weidner, VidalBralo, and Zhang), phenotypic age (PhenoAge, GrimAge, GrimAge2, and DunedinPoAm), and other (HorvathTelo, which estimates telomere length, and YangCell, which estimates cellular division rate). So that all age measures were on the same scale (years), we transformed DunedinPoAm from a rate of aging to years by multiplying it by participants’ reported age. Additionally, we reversed the direction of HorvathTelo in regression models to calculate associations per unit decrease in telomere length rather than an increase.

We calculated age deviation for measures of chronological and phenotypic age by regressing the DNA methylation measure against calendar age and extracting the model residuals (**Figure 1**). An age deviation value greater than 0 indicates that an individual’s epigenetic age was older than their expected calendar age, which we defined as age acceleration. An age deviation value less than 0 indicates that the epigenetic age was younger. HorvathTelo and YangCell were z-score standardized (mean=0 and standard deviation=1). As a secondary approach, we also z-score standardized all age deviation measures for unified interpretation across clock types as a one-standard deviation change in the measure. All transformations were conducted in the full DNA methylation subset (N=2,532), prior to further exclusions for missing outcome or covariate data.

**Figure 1.**
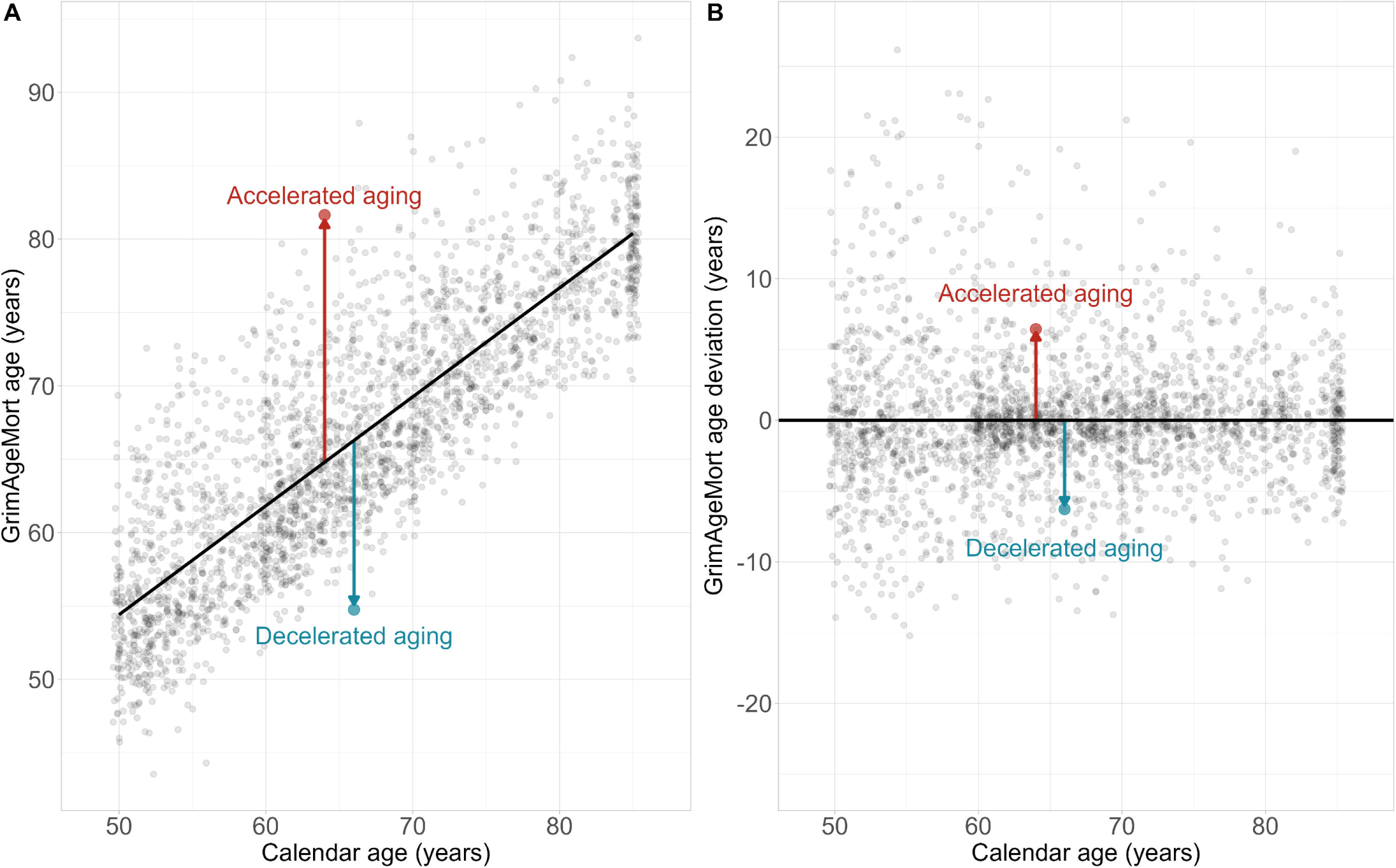
Scatterplots depicting an example calculation of DNA methylation age deviation with GrimAgeMort among NHANES participants, 1999-2002 (N=2,532). Panel A shows calendar age (x) versus GrimAgeMort age (y), with a solid black line indicating the line of best fit. Age deviation is calculated as the residuals of the regression model y=x. Panel B shows calendar age (x) versus GrimAgeMort age deviation (y), with a solid black line plotted at age deviation=0. The red and blue arrows indicate which sides of the dividing lines represent participants with accelerated aging and decelerated aging, respectively.

### Cognitive measures

Cognitive function was evaluated in participants aged 60 years and older with the Digit Symbol Substitution Test (DSST).^35^ The DSST is based on a module from the Wechsler Adult Intelligence Scale, Third Edition (WAIS-III).^36^ It involves a key of symbols, such as arrows and squares, that are paired with numbers. Participants are given a form with rows of numbers and must draw the matching symbols under each number. In NHANES, participants were given two minutes and scored on the total number of correct symbols drawn in that time. The maximum possible score was 133. We evaluated cognitive function as the continuous DSST score and as a binary mild cognitive impairment (MCI) variable, dichotomized at the weighted 25^th^ percentile. DSST scores below this cutoff were considered MCI.

### Covariate measures

Participants’ calendar age, sex, race and ethnicity, level of education, and tobacco use history were collected during the NHANES interview phase. To reduce identifiability, adults aged 85 or older were top-coded at 85. NHANES categorized sex as male or female and race and ethnicity as Mexican American, Other Hispanic, Non-Hispanic White, Non-Hispanic Black, or Other Race/Multiracial. Educational level was categorized as less than high school, high school/GED equivalent or some college, or completed college. Smoking status was categorized as never smokers (smoked fewer than 100 cigarettes in lifetime), former smokers (had smoked 100+ cigarettes and stopped), or current smokers (reported current tobacco smoking).

Body mass index (BMI) and serum cotinine were measured in the mobile examination center. Participants’ height (m) and weight (kg) were measured and used to calculate BMI (kg/m^2^). Serum cotinine (ng/mL) was measured from participant blood samples using isotope dilution-high performance liquid chromatography / atmospheric pressure chemical ionization tandem mass spectrometry.^37^ Cotinine was log2-transformed and z-score standardized for regression models.

NHANES additionally used the DNA methylation data to estimate cell type proportions with a regression calibration algorithm.^23^ Six cell types (B-lymphocytes, CD4+ T-cells, CD8+ T-cells, monocytes, natural killer cells, and neutrophils) were estimated and released in the DNA methylation public dataset. We converted the estimates from proportions to percentages.

### Statistical analyses

Analyses were conducted in R version 4.4.0 and RStudio.^38,39^ Code to produce all analyses is available (https://github.com/bakulskilab/NHANES_DNAm_cognition). Participants were included in the analysis if they participated in both the DNA methylation subsample and the DSST and had complete covariate data (age, sex, race/ethnicity, education, smoking, cotinine, and BMI). We visualized participant inclusion using a flow chart and compared the distributions of the included and excluded samples.

All analyses were survey-weighted using the ‘survey’ package to account for the NHANES sampling design, unless otherwise noted.^40^ We first calculated descriptive statistics for all DNA methylation measures, DSST scores, and covariates as means and standard deviations for continuous variables or proportions for categorical variables. We compared descriptive statistics of included versus excluded NHANES participants. Within the included sample, we compared participants by cognitive status (MCI versus no MCI) and by age acceleration status (accelerated aging, residuals >0, versus non-accelerated aging, residuals <0) using GrimAgeMort.

To evaluate the associations between epigenetic age acceleration and cognitive function, we ran a separate weighted linear regression model between each DNA methylation measure and DSST, adjusted for age, sex, race and ethnicity, education, smoking, cotinine, and BMI. For age deviation measures, we ran models both for an additional one year and one standard deviation of accelerated aging, while the telomere and mitotic measures were run only per one standard deviation change. We accounted for multiple testing by adjusting p-values with the Benjamini-Hochberg false discovery rate (FDR) method and used a significance threshold of adjusted p<0.05.^41^ We visualized regression results per one standard deviation change in the DNA methylation measures in a forest plot. For comparison, we calculated the association between calendar age and DSST, accounting for sex, race and ethnicity, education, smoking, cotinine, and BMI, and reported the relationship between each one-year increase in calendar age with DSST.

### Sensitivity analyses

We conducted four sensitivity analyses to further investigate these associations. First, we evaluated the binary MCI outcome in place of the continuous DSST score. We ran modified Poisson regression models for the associations between each DNA methylation measure and binary cognitive function, adjusted for the same set of covariates.

Second, since cell type composition can be related to DNA methylation as well as cognition, we considered including estimated DNA methylation cell type percentages as an additional confounder in two approaches.^42^ We conducted principal component analysis (PCA) on the six cell types estimated in the DNA methylation dataset: CD8+ T cells, CD4+ T cells, natural killer cells, B cells, monocytes, and neutrophils. We adjusted models further for PC1 and PC2, which accounted for 63.9% of variance (**Supplemental Figure 1**). Separately, we adjusted models for five of the six cell types, excluding neutrophils, which represented the highest proportion.

Third, in NHANES, age was top-coded at 85 years to reduce identifiability. Thus, among participants top-coded at 85, our study may overestimate their DNA methylation age deviation. We restricted our analyses to participants below age 85 and re-ran the analyses.

Fourth, we tested for interaction between DNA methylation measures and sex, education, and race and ethnicity. For each of these variables of interest, we added an interaction term between the DNA methylation measure and the variable to the regression models. We presented the associations between DNA methylation measures and DSST score for each category of the interactor (male, female; less than high school, high school/GED or some college, completed college; Non-Hispanic White, Non-Hispanic Black, Mexican American, Other Hispanic, or Other Race/Multiracial).

## Results

### Study sample descriptive statistics

Of the 2,532 participants with complete DNA methylation data, 1,550 also had a DSST score. Participants were further excluded if they were missing any covariate data, for an analytic sample size of N=1,463 (**Figure 2**). In comparison to the included sample, excluded participants in the DNA methylation subset that were also eligible for the DSST were older, had lower DSST scores, were less likely to be Non-Hispanic White, and were less likely to have completed high school (**Supplemental Table 1**). Most DNA methylation age acceleration measures were not significantly different between the included and excluded groups; however, included participants had lower PhenoAge, DunedinPoAm, and HorvathTelo measures and higher YangCell measures.

**Figure 2.**
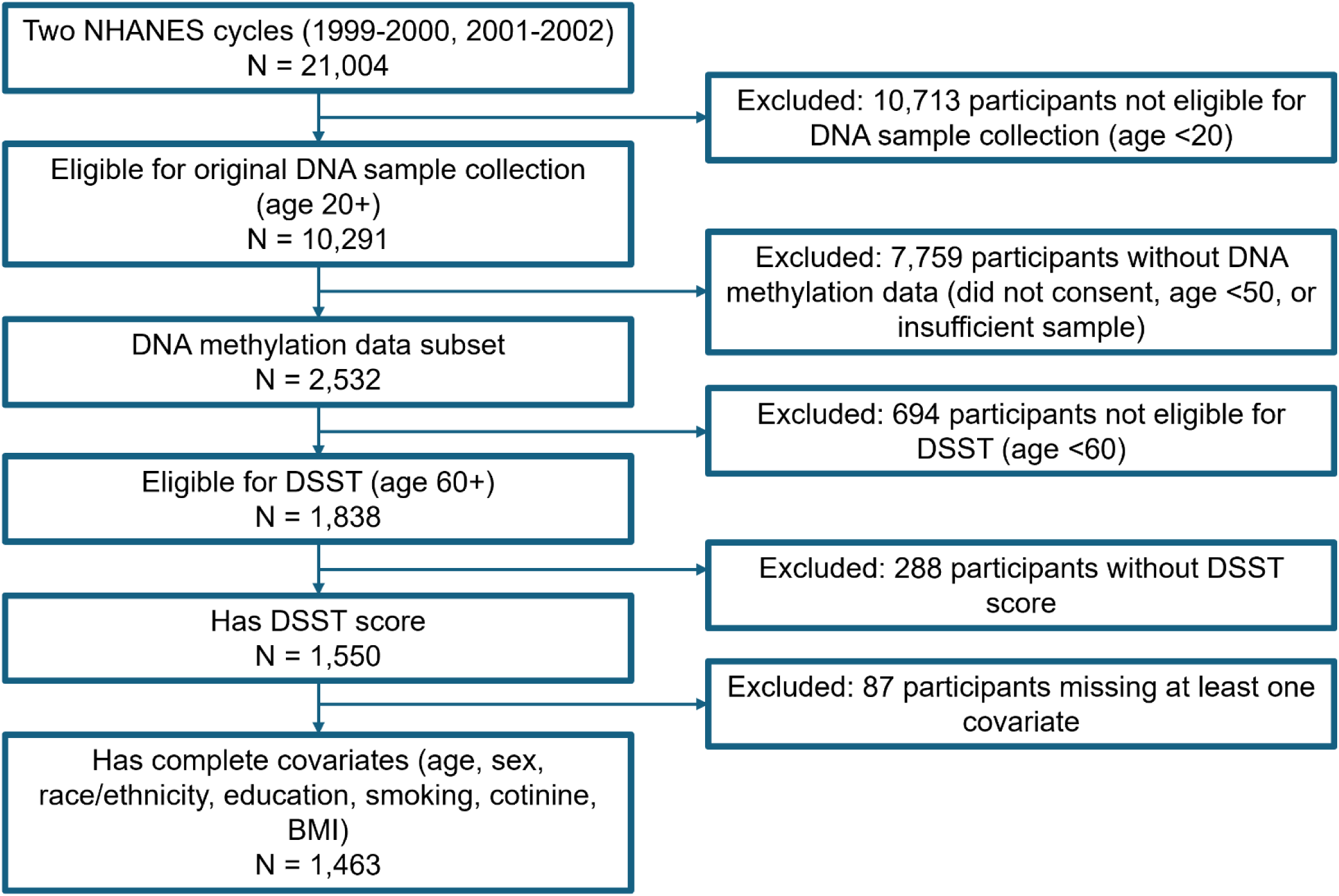
Flowchart of inclusion/exclusion criteria for participants in the National Health and Nutrition Examination Survey (NHANES), 1999-2002.

Included participants (N=1,463) were on average 70.5 years, 56.9% female, and 82.7% Non-Hispanic White (**Table 1**). The average DSST score was 46.9 and N=581 participants were categorized as having MCI (DSST<35, weighted 25^th^ percentile). Participants with MCI were less likely to complete high school or college and had higher age acceleration measured by PhenoAge, GrimAgeMort, and GrimAge2Mort clocks. Participants with accelerated GrimAgeMort had lower DSST scores, were more likely to be male, and were more likely to be former or current smokers.

**Table 1.** Weighted descriptive statistics of included participants in the National Health and Nutrition Examination Survey (NHANES) 1999-2002, stratified by cognitive impairment status and GrimAgeMort acceleration.

| Variable | Overall<br>(N = 1,463)<sup>1</sup> | Mild cognitive<br>impairment<br>(N = 581)<sup>1</sup> | No cognitive<br>impairment<br>(N = 882)<sup>1</sup> | p-value<sup>2</sup> | Non-accelerated<br>GrimAgeMort<br>(N = 813)<sup>1</sup> | Accelerated<br>GrimAgeMort<br>(N = 650)<sup>1</sup> | p-value<sup>2</sup> |
| --- | --- | --- | --- | --- | --- | --- | --- |
| **Digit Symbol Substitution Test score** | 46.9 (17.6) | 23.7 (8.8) | 54.4 (12.5) | <0.001 | 49.1 (18.2) | 43.8 (16.2) | <0.001 |
| **Age (years)** | 70.5 (7.3) | 73.5 (7.8) | 69.6 (6.9) | <0.001 | 70.3 (7.3) | 70.8 (7.2) | 0.4 |
| **Sex** |  |  |  | >0.9 |  |  | <0.001 |
| Male | 43.1% | 43.2% | 43.0% |  | 32.9% | 57.7% |  |
| Female | 56.9% | 56.8% | 57.0% |  | 67.1% | 42.3% |  |
| **Race/ethnicity** |  |  |  | <0.001 |  |  | 0.02 |
| Mexican American | 2.8% | 6.2% | 1.7% |  | 2.8% | 2.9% |  |
| Other Hispanic | 4.7% | 9.5% | 3.2% |  | 5.8% | 3.1% |  |
| Non-Hispanic White | 82.7% | 65.4% | 88.2% |  | 82.3% | 83.2% |  |
| Non-Hispanic Black | 7.1% | 15.9% | 4.3% |  | 6.0% | 8.7% |  |
| Other Race | 2.7% | 3.0% | 2.6% |  | 3.1% | 2.1% |  |
| **Education** |  |  |  | <0.001 |  |  | 0.049 |
| Less than high school | 28.4% | 60.4% | 18.2% |  | 25.3% | 32.9% |  |
| High school or some college | 51.1% | 34.0% | 56.6% |  | 51.4% | 50.8% |  |
| College or above | 20.4% | 5.6% | 25.2% |  | 23.3% | 16.3% |  |
| **Poverty-income ratio** | 2.9 (1.6) | 1.9 (1.3) | 3.2 (1.5) | <0.001 | 3.0 (1.6) | 2.7 (1.6) | 0.02 |
| Missing | 162 | 65 | 97 |  | 92 | 70 |  |
| **Smoking status** |  |  |  | 0.5 |  |  | <0.001 |
| Never smoker | 45.7% | 48.9% | 44.7% |  | 65.6% | 17.2% |  |
| Former smoker | 42.5% | 39.0% | 43.6% |  | 32.5% | 56.9% |  |
| Current smoker | 11.7% | 12.1% | 11.6% |  | 1.9% | 25.9% |  |
| **Serum cotinine (ng/mL)** | 29.6 (92.0) | 42.1 (126.1) | 25.7 (77.6) | 0.07 | 10.1 (69.6) | 57.8 (111.0) | <0.001 |
| **Body mass index (kg/m<sup>2</sup>)** | 28.13 (5.45) | 27.79 (5.63) | 28.24 (5.39) | 0.2 | 27.8 (5.1) | 28.6 (5.9) | 0.02 |
| **Chronological age deviation** |  |  |  |  |  |  |  |
| **HorvathAge** | -0.08 (9.2) | -0.02 (9.3) | -0.1 (9.2) | >0.9 | -1.6 (9.4) | 2.1 (8.5) | <0.001 |
| **HannumAge** | -0.2 (6.9) | 0.3 (6.9) | -0.3 (6.9) | 0.3 | -1.6 (7.0) | 2.0 (6.2) | <0.001 |
| **SkinBloodAge** | 0.03 (6.0) | 0.06 (6.1) | 0.01 (6.0) | >0.9 | -0.8 (6.5) | 1.2 (5.1) | <0.001 |
| **LinAge** | -0.3 (10.5) | -0.3 (8.9) | -0.3 (11.0) | >0.9 | -2.0 (10.3) | 2.2 (10.3) | <0.001 |
| **WeidnerAge** | -0.4 (12.2) | 0.1 (11.4) | -0.6 (12.4) | 0.6 | -1.7 (11.4) | 1.4 (12.9) | 0.02 |
| **VidalBraloAge** | 0.02 (7.3) | -0.2 (6.5) | 0.1 (7.5) | 0.6 | -1.5 (6.7) | 2.2 (7.5) | <0.001 |
| **ZhangAge** | 0.05 (2.2) | 0.02 (2.4) | 0.06 (2.2) | 0.8 | -0.2 (2.5) | 0.4 (1.8) | <0.001 |
| **Phenotypic age deviation** |  |  |  |  |  |  |  |
| **PhenoAge** | -0.5 (8.5) | 1.1 (8.2) | -1.0 (8.5) | 0.01 | -2.8 (8.5) | 2.8 (7.3) | <0.001 |
| **GrimAgeMort** | -0.3 (6.0) | 0.4 (5.1) | -0.6 (6.3) | 0.006 | -4.2 (3.0) | 5.3 (4.8) | <0.001 |
| **GrimAge2Mort** | -0.4 (6.5) | 0.7 (5.4) | -0.8 (6.8) | <0.001 | -4.5 (3.7) | 5.4 (5.1) | <0.001 |
| **DunedinPoAm** | -0.4 (8.4) | 0.1 (8.4) | -0.6 (8.5) | 0.2 | -4.1 (6.9) | 4.8 (7.8) | <0.001 |
| **Other DNA methylation measures** |  |  |  |  |  |  |  |
| **HorvathTelo<sup>3</sup>** | -0.3 (0.9) | -0.5 (1.0) | -0.3 (0.9) | 0.01 | -0.07 (0.8) | -0.7 (0.9) | <0.001 |
| **YangCell<sup>3</sup>** | -0.1 (0.9) | 0.3 (1.1) | -0.2 (0.9) | <0.001 | -0.2 (0.9) | -0.1 (1.0) | 0.09 |
| **Estimated cell type percentages** |  |  |  |  |  |  |  |
| **CD8+ T cell percentage** | 8.6 (4.1) | 9.3 (4.6) | 8.4 (3.9) | 0.02 | 8.7 (4.1) | 8.5 (4.2) | 0.4 |
| **CD4+ T cell percentage** | 16.3 (6.6) | 15.5 (6.4) | 16.5 (6.6) | 0.02 | 17.3 (6.6) | 14.7 (6.2) | <0.001 |
| **Natural killer cell percentage** | 6.1 (2.7) | 6.5 (2.8) | 5.9 (2.6) | 0.03 | 6.2 (2.7) | 5.8 (2.6) | 0.02 |
| **B cell percentage** | 6.1 (3.2) | 5.9 (2.9) | 6.1 (3.3) | 0.3 | 6.3 (2.9) | 5.7 (3.6) | 0.02 |
| **Monocyte percentage** | 8.0 (2.4) | 8.1 (2.3) | 8.0 (2.4) | 0.6 | 7.9 (2.3) | 8.2 (2.4) | 0.2 |
| **Neutrophil percentage** | 60.0 (10.5) | 59.9 (10.6) | 60.1 (10.5) | 0.8 | 58.7 (10.2) | 62.0 (10.8) | <0.001 |

<sup>1</sup>Mean (SD); %.
<sup>2</sup>Design-based t-test; Pearson's X<sup>2</sup>: Rao & Scott adjustment.
<sup>3</sup>Measures are z-score standardized.
Mild cognitive impairment is defined as DSST score less than the weighted 25<sup>th</sup> percentile. Accelerated GrimAgeMort is defined as GrimAgeMort age acceleration greater than 0.

### Adjusted associations between DNA methylation measures and cognition

In covariate-adjusted linear regression models, for all DNA methylation measures, the direction of association was that higher DNA methylation measures reflected lower DSST scores (**Figure 3**). **Table 2** depicts the regression results for both a one-year and one-standard deviation change in the DNA methylation measures. After accounting for multiple comparisons, 10 of 13 DNA methylation measures were associated with DSST scores, and these included measures from all four aging marker types (chronological, phenotypic, telomere, and mitotic). Among the chronological epigenetic clocks, one year of additional ZhangAge accelerated aging was associated with –0.61 points lower DSST score (95% CI: –1.12, –0.09). Among the phenotypic clocks, one year increase in GrimAge2Mort accelerated aging was associated with –0.41 points lower DSST score (95% CI: – 0.61, –0.21). For comparison, each one-year increase in calendar age was associated with –0.89 points lower DSST (95% CI: –1.00, –0.77).

**Figure 3.**
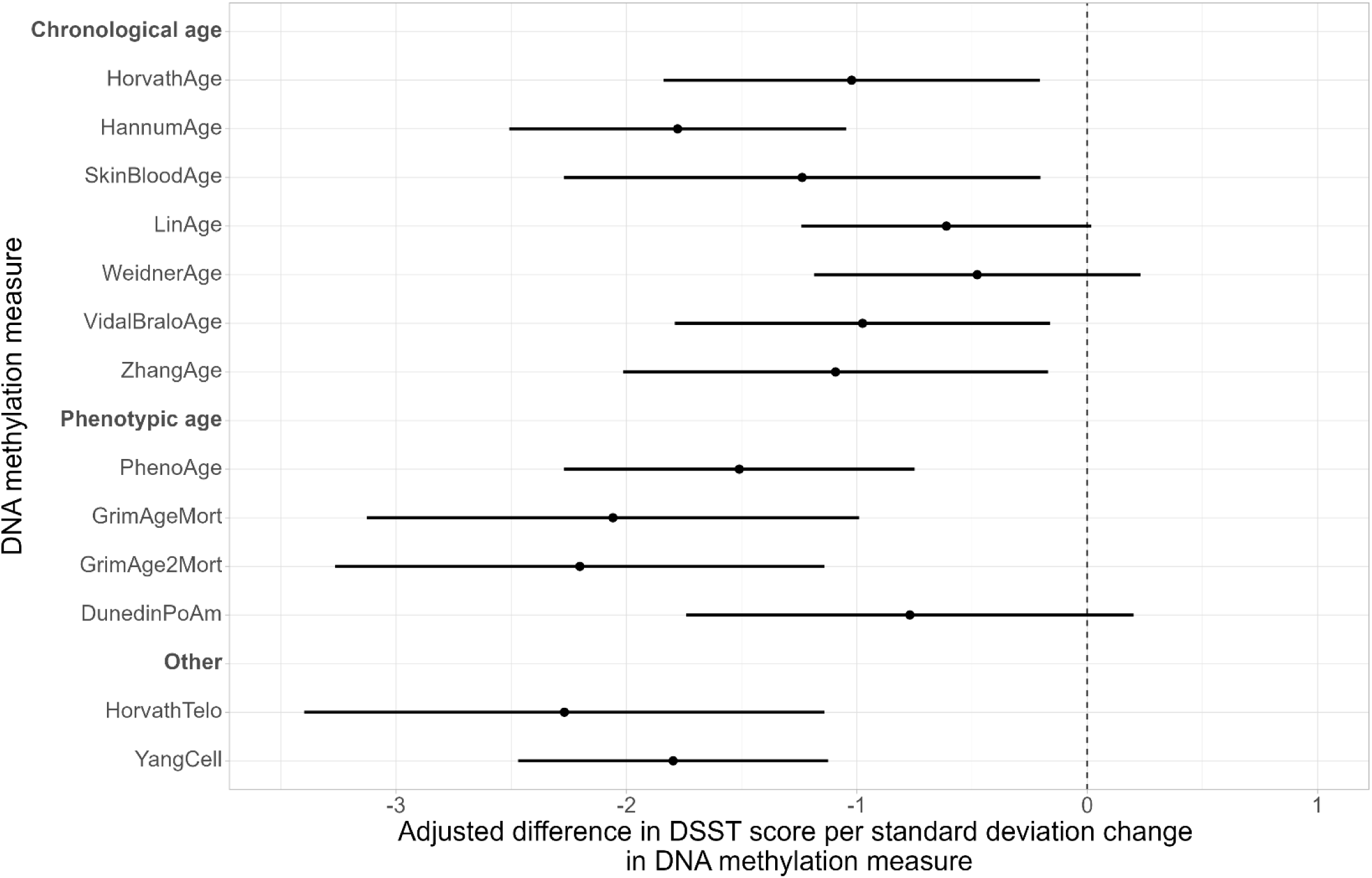
Forest plot of regression coefficients and 95% confidence intervals for the adjusted associations between z-score standardized DNA methylation measures and Digit Symbol Substitution Test (DSST) scores.

**Table 2.**
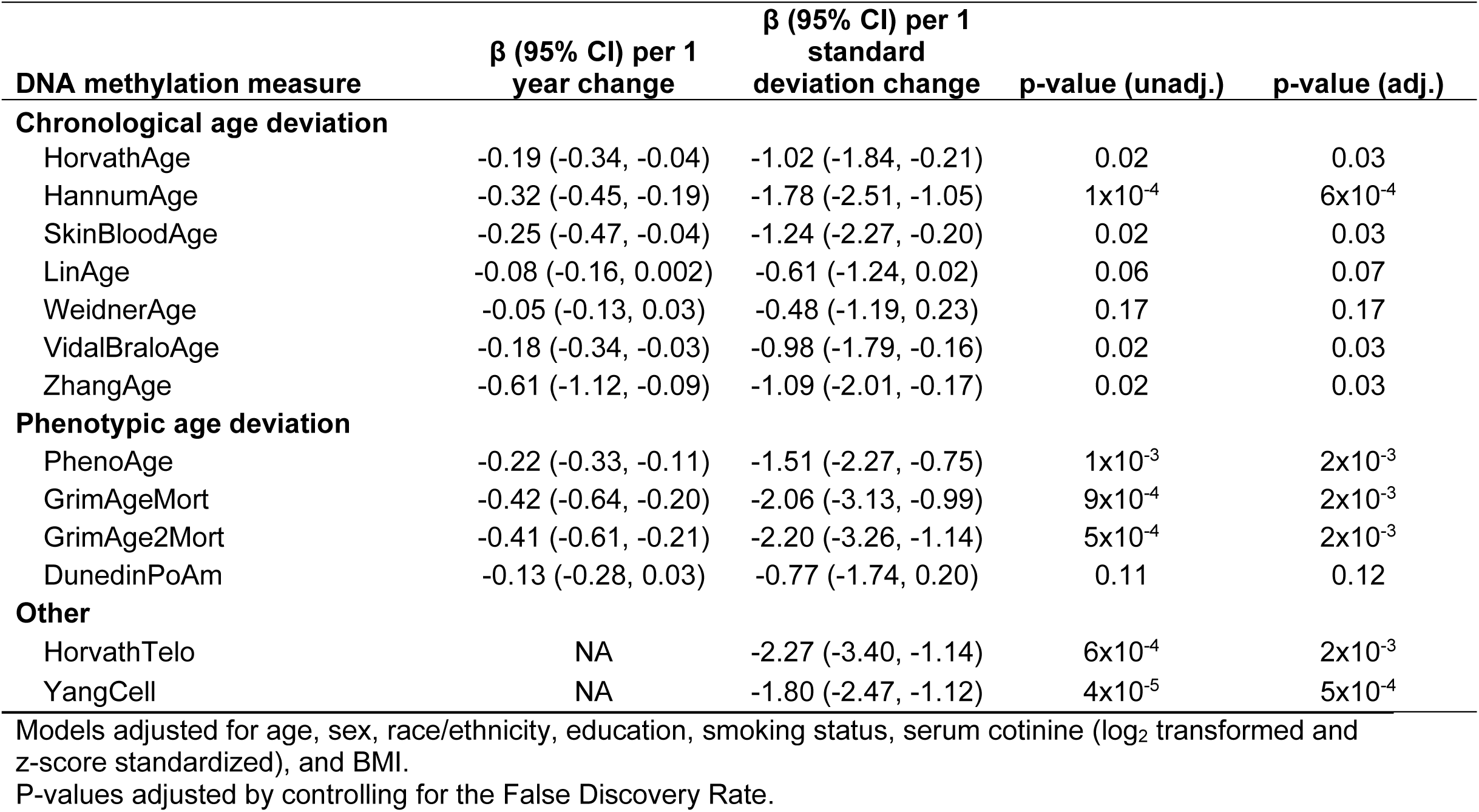
Summary of survey-weighted linear regression models evaluating the association between DNA methylation measures and Digit Symbol Substitution Test scores in NHANES, 1999-2002 (N = 1,463).

For DNA methylation estimated telomere length, a one standard-deviation decrease in HorvathTelo was associated with –2.27 points lower DSST score (95% CI: –3.40, –1.14). For cellular division, a one standard-deviation increase in the mitotic clock (YangCell) was associated with –1.80 points lower DSST score (95% CI: –2.47, –1.12). LinAge and WeidnerAge (chronological) and DunedinPoAm (phenotypic) were the three measures not associated with DSST after multiple comparison adjustment.

### Results of sensitivity analyses

Although DNA methylation age acceleration was associated with continuous DSST scores, no associations with the binary MCI outcome were found after adjustment for multiple comparisons (**Supplemental Table 2**). When adjusting models for cell type PCA and for cell type percentages, we saw similar results to the main models (**Supplemental Table 3**). The magnitudes of association were consistent, except for the YangCell clock (main model β=–1.80; cell type PCA adjusted β=–2.52; cell percentage adjusted β=–2.43). In models restricted to participants aged less than 85 (N=1,393), the magnitudes of association were again consistent with the overall analyses, although three clocks (SkinBloodAge, VidalBraloAge, and ZhangAge) were no longer significant after multiple testing adjustment (**Supplemental Table 4**).

### Effect modification by sex, education, and race/ethnicity

We identified significant statistical interaction by sex with SkinBloodAge age deviation (p=0.02); more pronounced negative associations with DSST were found in male compared to female participants (**Supplemental Table 5**). Interactions by education status and race/ethnicity were not significant, although we noted that the associations between DNA methylation and DSST tended to only be significant among Non-Hispanic White participants (**Supplemental Tables 6 and 7**).

## Discussion

In a United States nationally representative sample of adults ages 60+, we observed that ten biomarkers of epigenetic age estimated from venous blood are cross-sectionally associated with cognition. These associations included DNA methylation clocks of chronological age (HorvathAge, HannumAge, SkinBloodAge, VidalBraloAge, and ZhangAge), phenotypic age (PhenoAge, GrimAgeMort, and GrimAge2Mort), telomere length (HorvathTelo), and cellular division rate (YangCell), showing consistency across hallmarks of aging. For example, each one-year increase in GrimAge2Mort aging was associated with –0.41 points lower DSST score (95% CI: –0.61, –0.21), which is approximately half of the magnitude of the association between calendar age and DSST score. While many circulating biomarkers are currently available for Alzheimer’s disease (e.g. phosphotau217, beta amyloid 42:40 ratio),^4^ indicators of earlier susceptibility, as estimated here by lower general cognition, are less available. These findings suggest DNA methylation age estimates may be markers of cognitive status among older adults, and future studies can investigate temporality and longitudinal outcomes.

These findings are consistent with the literature for phenotypic clocks. For example, across three United Kingdom studies of adults ages 45-87 (Medical Research Council National Survey of Health and Development (n=2,047), National Child Development Study (n=240), and TwinsUK (n=120)), the phenotypic clocks, GrimAge and PhenoAge, were associated (p<0.05) with lower episodic memory and mental speed, which is similar to our results.^17^ However, the United Kingdom studies did not observe associations for the chronological clocks, Horvath and Hannum, with episodic memory or mental speed, in contrast with our findings.^17^ Our findings build on existing research on phenotypic clocks with cognition, and provide support for relationships with chronological, telomere, and mitotic clocks as well.

Previous research has presented relationships between DNA methylation age deviation and cognition at a subsequent visit. Among 486 monozygtic twins (243 pairs) in the Middle Aged Danish Twin Study, accelerated DNA methylation age (measured in 1998) was not associated with cognition (measured between 2008 and 2011) after follow up (p=0.39 for Horvath, p=0.82 for Hannum) in paired analyses, though this study did not examine additional DNA methylation age measures.^19^ In the Adult Health and Behavior Study, 48 adults mean age 44.7 at baseline had two measures of DNA methylation age and cognition 16 years apart.^20^ They observed persons with cognitive decline had older GrimAge and DunedinPoAm than those that maintained their cognition levels over follow up.^20^ Similar to this study, we observed GrimAge was associated with cognition, though we did not observe an association with DunedinPoAm. Due to the design of NHANES, we were limited to a single DNA methylation measure and a single cross-sectional cognitive measure. Studies such as in the Adult Health and Behavior Study with repeated measures of DNA methylation in the same participants are rare and future studies can expand on this approach.

Additional studies have examined the relationship between DNA methylation aging biomarkers with cognitive outcomes that are further along in the disease process. For example, among 3,724 adults over age 50 in the United States Health and Retirement Study, a one-year increase in GrimAge acceleration was associated with 1.6 times greater odds of cognitive impairment cross-sectionally, relative to normal cognition (95% CI: 1.3-2.1).^43^ These findings are in the same direction as our study, though we did not have a clinically validated cognitive impairment measure, as in the Health and Retirement Study. This study also observed an additive interaction by education status, which we did not.^43^ In a prospective analysis also in the Health and Retirement Study, among 2,713 adults, a one-year increase in baseline GrimAge acceleration was associated with 1.08 times higher odds of cognitive decline (95% CI: 1.05, 1.11), defined as a transition from either normal cognition to cognitive impairment or dementia, or a transition from impairment to dementia, over two years of follow up.^44^ Our study examining subclinical cognition complements existing research on discrete impairment and dementia outcomes.

Our study has several strengths, which advance the literature. First, the use of NHANES with survey weights, as we have done in this study, results in findings that are nationally representative of the non-institutionalized population of the United States. This greatly improves the generalizability of the observations. Second, this is a relatively large sample size (n=1,463) of older adults (ages 60+), which improves the rigor of research in this understudied age group. Third, we used 13 DNA methylation aging biomarkers, which span markers of chronological age, phenotypic age, telomere length, and cell senescence. This allowed us to assess multiple dimensions of biologic aging, and while accounting for these multiple comparisons, we observed relatively consistent findings across all marker types. Fourth, we used DSST, which is derived from the Wechsler Adult Intelligence Scale, and provides a validated measure of global cognitive function. Fifth, we performed several sensitivity analyses, including modeling a binary outcome, additional covariate adjustment, age restriction, and supplemental analyses of effect modification, which improve the study completeness.

Our study has several limitations that point to directions for future study. First, this is a cross-sectional study. Blood collection for DNA methylation measures and cognition tests was performed at the same study visit. We cannot make inferences about temporality or causality, and future studies may be able to build on this work to analyze prospective or repeated measures. Second, while our study was nationally representative at the time of study (1999-2002), it does not reflect the current demographics of the United States. Our sample was predominantly non-Hispanic White, and it may have been underpowered to detect effect modification across social groups. Additionally, many of these epigenetic clocks in NHANES vary in correlation with calendar age by race and ethnicity, with lower performance among non-Hispanic Black and Mexican American participants, which may limit the interpretation of our effect modification results.^45^ Third, to reduce identifiability, NHANES top codes ages greater than 85 to 85. While we performed a sensitivity analysis stratified to ages less than 85 and showed our findings were consistent with the full sample, this design limits our ability to make inferences about the association between DNA methylation age and cognition beyond age 85.

In summary, we observed DNA methylation age estimates are consistently and robustly associated with cross-sectional cognition among older adults in a United States nationally representative sample. These findings are consistent with existing studies on phenotypic clocks and cognition, and also demonstrate associations for chronological, telomere, and mitotic clocks with cognition. This research suggests DNA methylation age measures in blood may be biomarkers for susceptibility for lower cognition, which complements existing biomarkers for clinical diagnoses, such as Alzheimer’s disease. Identifying susceptibility biomarkers is a priority in the field and will allow for early detection and risk stratification. Future studies will assess the temporality and prediction capability of these markers.

## Data Availability

NHANES datasets used in this analysis are publicly available online from the NCHS (https://www.n.cdc.gov/nchs/nhanes/default.aspx).

## Acknowledgements

We thank the participants of NHANES and the CDC staff who collected and provided the data. This analysis was supported by the National Institutes of Health, specifically the National Institute on Aging under R01 AG070897, P30 AG072931, and R01 AG067592 and the National Institute on Minority Health and Health Disparities under R01 MD011721.

## Data availability statement

NHANES datasets are publicly available online from the NCHS (https://www.n.cdc.gov/nchs/nhanes/default.aspx). The code to perform this analysis is available on GitHub (https://github.com/bakulskilab/NHANES_DNAm_cognition).

