## Supplement for "DNA methylation age deviation and cognitive status among older adults in the US, NHANES 1999-2002"

### Supplemental figures and tables

**Supplemental Figure 1.** Plots depicting information on principal component analysis (PCA) of predicted cell type percentages among included participants (N=1,463). Panel A shows the percentage of variance explained by each principal component. Panel B shows a scatterplot of loadings for PC1 versus PC2.

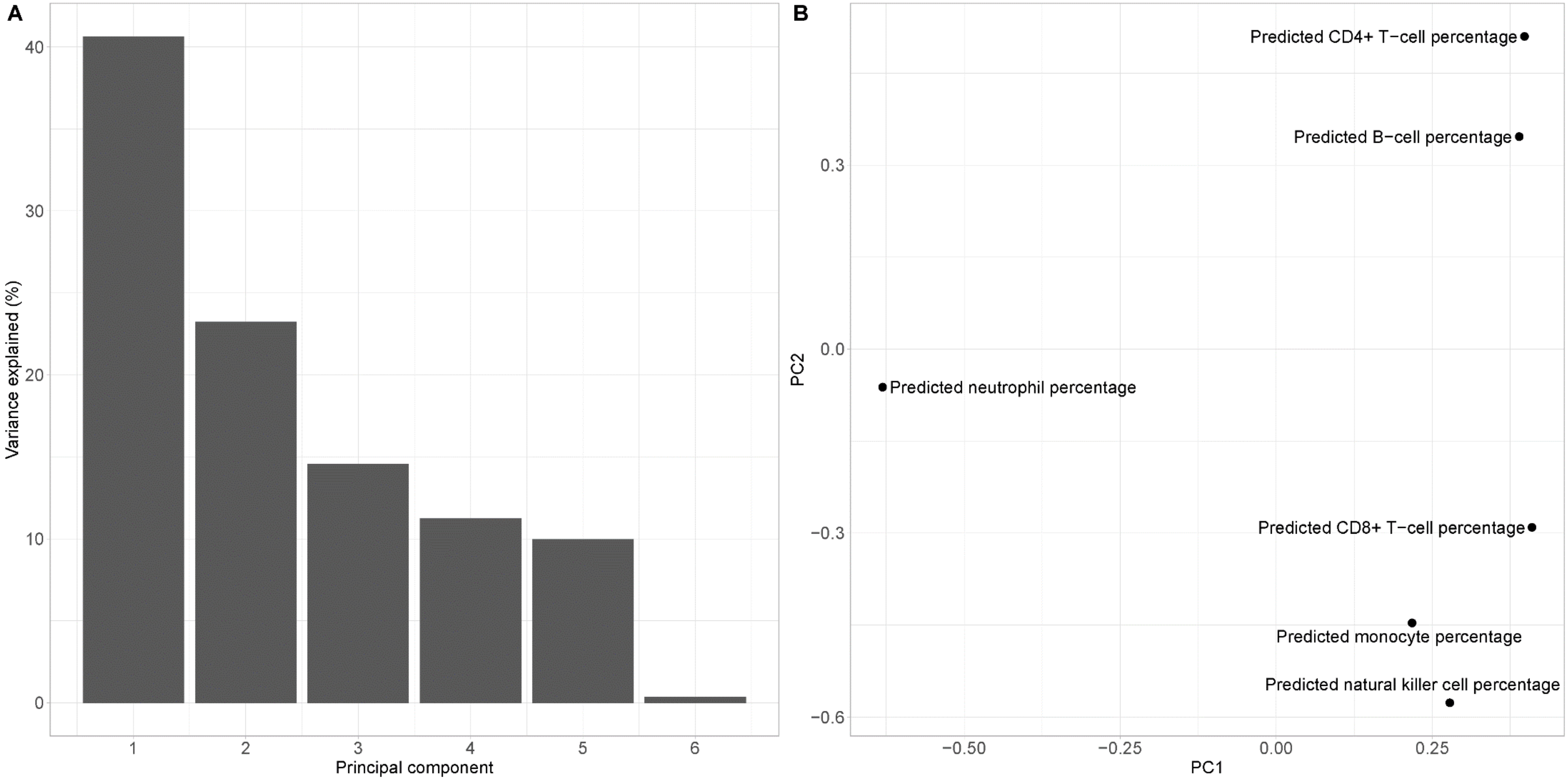

**Supplemental Table 1.** Weighted descriptive statistics of included vs. excluded participants in the National Health and Nutrition Examination Survey (NHANES), 1999-2002; subset of participants aged 60 years and older with complete DNA methylation data (N = 1,838).

| Variable | Excluded (N = 375)[1] | Included (N = 1,463)[1] | p-value[2] |
|---|---|---|---|
| **Digit Symbol Substitution Test score** | 39.7 (19.1) | 46.9 (17.6) | 0.01 |
| Missing | 288 | 0 | |
| **Age (years)** | 73.9 (8.3) | 70.5 (7.3) | <0.001 |
| **Sex** | | | 0.6 |
| Male | 44.8% | 43.1% | |
| Female | 55.2% | 56.9% | |
| **Race/ethnicity** | | | <0.001 |
| Mexican American | 4.5% | 2.8% | |
| Other Hispanic | 7.3% | 4.7% | |
| Non-Hispanic White | 70.8% | 82.7% | |
| Non-Hispanic Black | 14.5% | 7.1% | |
| Other Race | 2.9% | 2.7% | |
| **Education** | | | 0.006 |
| Less than high school | 44.4% | 28.4% | |
| High school or some college | 44.4% | 51.1% | |
| College or above | 11.2% | 20.4% | |
| Missing | 4 | 0 | |
| **Poverty-income ratio** | 2.3 (1.4) | 2.9 (1.6) | <0.001 |
| Missing | 57 | 162 | |
| **Smoking status** | | | 0.4 |
| Never smoker | 49.8% | 45.7% | |
| Former smoker | 41.3% | 42.5% | |
| Current smoker | 8.9% | 11.7% | |
| Missing | 3 | 0 | |
| **Serum cotinine (ng/mL)** | 45.0 (122.8) | 29.7 (92.0) | 0.046 |
| Missing | 50 | 0 | |
| **Body mass index (kg/m$^2$)** | 28.1 (6.0) | 28.1 (5.5) | 0.6 |
| Missing | 104 | 0 | |
| **Chronological age deviation** | | | |
| **HorvathAge** | 0.8 (5.8) | -0.1 (6.9) | 0.1 |
| **HannumAge** | 0.7 (7.2) | -0.2 (6.9) | 0.3 |
| **SkinBloodAge** | 0.5 (6.2) | 0.03 (6.0) | 0.5 |
| **LinAge** | 0.5 (9.0) | -0.3 (10.5) | 0.5 |
| **WeidnerAge** | 1.5 (12.1) | -0.4 (12.2) | 0.08 |
| **VidalBraloAge** | 0.7 (7.8) | 0.02 (7.3) | 0.4 |

| Variable | Excluded (N = 375)[1] | Included (N = 1,463)[1] | p-value[2] |
|---|---|---|---|
| **ZhangAge** | 0.3 (1.9) | 0.05 (2.2) | 0.3 |
| **Phenotypic age deviation** | | | |
| **PhenoAge** | 1.6 (7.9) | -0.5 (8.5) | 0.04 |
| **GrimAgeMort** | 0.1 (4.6) | -0.3 (6.0) | 0.5 |
| **GrimAge2Mort** | 0.5 (5.2) | -0.4 (6.5) | 0.1 |
| **DunedinPoAm** | 1.3 (7.2) | -0.4 (8.4) | 0.003 |
| **Other DNA methylation measures** | | | |
| **HorvathTelo[3]** | -0.5 (0.9) | -0.3 (0.9) | 0.03 |
| **YangCell[3]** | 0.1 (1.1) | -0.1 (1.0) | 0.007 |
| **Estimated cell type percentages** | | | |
| **CD8+ T cell percentage** | 8.4 (4.3) | 8.6 (4.1) | 0.7 |
| **CD4+ T cell percentage** | 15.3 (7.0) | 16.3 (6.6) | 0.2 |
| **Natural killer cell percentage** | 6.1 (2.5) | 6.1 (2.7) | 0.8 |
| **B cell percentage** | 6.1 (3.7) | 6.1 (3.2) | >0.9 |
| **Monocyte percentage** | 8.5 (2.8) | 8.0 (2.4) | 0.08 |
| **Neutrophil percentage** | 60.6 (12.0) | 60.0 (10.6) | 0.7 |

[1]Mean (SD); %; unweighted N missing.

[2]Design-based t-test; Pearson's $X^2$: Rao & Scott adjustment.

[3]Measures are z-score standardized.

**Supplemental Table 2.** Summary of survey-weighted modified Poisson regression models evaluating the association between DNA methylation measures and mild cognitive impairment in NHANES, 1999-2002 (N = 1,463).

| DNA methylation measure | Risk ratio (95% CI) | p-value (unadj.) | p-value (adj.) |
|---|---|---|---|
| **Chronological age deviation** | | | |
| HorvathAge | 1.00 (0.99, 1.02) | 0.75 | 0.81 |
| HannumAge | 1.01 (0.99, 1.03) | 0.20 | 0.56 |
| SkinBloodAge | 1.00 (0.98, 1.03) | 0.69 | 0.81 |
| LinAge | 1.00 (0.99, 1.01) | 0.94 | 0.94 |
| WeidnerAge | 1.00 (0.99, 1.01) | 0.51 | 0.81 |
| VidalBraloAge | 1.00 (0.98, 1.01) | 0.70 | 0.81 |
| ZhangAge | 1.01 (0.95, 1.07) | 0.71 | 0.81 |
| **Phenotypic age deviation** | | | |
| PhenoAge | 1.02 (1.00, 1.04) | 0.06 | 0.26 |
| GrimAgeMort | 1.01 (0.99, 1.04) | 0.23 | 0.56 |
| GrimAge2Mort | 1.03 (1.00, 1.05) | 0.02 | 0.14 |
| DunedinPoAm | 1.00 (0.99, 1.02) | 0.68 | 0.81 |
| **Other** | | | |
| HorvathTelo | 1.07 (0.94, 1.22) | 0.26 | 0.56 |
| YangCell | 1.14 (1.03, 1.26) | 0.01 | 0.14 |

Models adjusted for age, sex, race/ethnicity, education, smoking status, serum cotinine ($log_2$ transformed and z-score standardized), and BMI.

Chronological and phenotypic age deviation models are per one-year change in age deviation. Other models are per one-standard deviation change.

P-values adjusted by controlling for the False Discovery Rate.

**Supplemental Table 3.** Summary of survey-weighted linear regression models evaluating the association between DNA methylation measures and Digit Symbol Substitution Test scores in NHANES, 1999-2002 (N = 1,463), adjusted further for cell type PCA or cell type percentages.

| DNA methylation measure | Main analysis | Adjusted for cell PCA | Adjusted for cell types |
|---|---|---|---|
| **Chronological age deviation** | | | |
| HorvathAge | -0.14 (-0.25, -0.03)* | -0.13 (-0.24, -0.01)* | -0.12 (-0.23, -0.006) |
| HannumAge | -0.32 (-0.45, -0.19)* | -0.31 (-0.45, -0.16)* | -0.32 (-0.48, -0.16)* |
| SkinBloodAge | -0.25 (-0.47, -0.04)* | -0.24 (-0.46, -0.02)* | -0.23 (-0.46, -0.01) |
| LinAge | -0.08 (-0.16, 0.00) | -0.07 (-0.15, 0.02) | -0.07 (-0.16, 0.02) |
| WeidnerAge | -0.05 (-0.13, 0.03) | -0.04 (-0.13, 0.04) | -0.04 (-0.13, 0.05) |
| VidalBraloAge | -0.18 (-0.34, -0.03)* | -0.19 (-0.34, -0.03)* | -0.18 (-0.34, -0.01) |
| ZhangAge | -0.61 (-1.12, -0.09)* | -0.56 (-1.08, -0.03) | -0.56 (-1.11, -0.01) |
| **Phenotypic age deviation** | | | |
| PhenoAge | -0.22 (-0.33, -0.11)* | -0.23 (-0.34, -0.11)* | -0.23 (-0.35, -0.11)* |
| GrimAgeMort | -0.42 (-0.64, -0.20)* | -0.43 (-0.63, -0.23)* | -0.42 (-0.62, -0.22)* |
| GrimAge2Mort | -0.41 (-0.61, -0.21)* | -0.44 (-0.63, -0.26)* | -0.44 (-0.62, -0.25)* |
| DunedinPoAm | -0.13 (-0.28, 0.03) | -0.13 (-0.29, 0.03) | -0.14 (-0.31, 0.03) |
| **Other** | | | |
| HorvathTelo | -2.27 (-3.40, -1.14)* | -2.18 (-3.54, -0.82)* | -2.06 (-3.49, -0.62)* |
| YangCell | -1.80 (-2.47, -1.12)* | -2.52 (-3.47, -1.57)* | -2.43 (-3.56, -1.30)* |

Main analysis models adjusted for age, sex, race/ethnicity, education, smoking status, serum cotinine, and BMI. PCA models adjusted for main analysis covariates, cell type PC1, and cell type PC2. Cell type models adjusted for main analysis covariates, CD8+, CD4+, natural killer, B cell, and monocyte percentages.

Chronological and phenotypic age deviation models are per one-year change in age deviation. Other models are per one-standard deviation change.

P-values adjusted by controlling for the False Discovery Rate (FDR).

* FDR adjusted $p < 0.05$.

**Supplemental Table 4.** Summary of survey-weighted linear regression models evaluating the association between DNA methylation measures and Digit Symbol Substitution Test (DSST) scores, NHANES 1999-2002, restricted to participants under age 85 (overall N = 1,393)

| DNA methylation measure | β (95% CI) | p-value (unadj) | p-value (adj) |
|---|---|---|---|
| **Chronological age deviation** | | | |
| HorvathAge | -0.14 (-0.26, -0.02) | 0.02 | 0.04 |
| HannumAge | -0.31 (-0.45, -0.17) | 0.0002 | 0.001 |
| SkinBloodAge | -0.23 (-0.45, -0.02) | 0.04 | 0.05 |
| LinAge | -0.08 (-0.16, 0.00) | 0.05 | 0.06 |
| WeidnerAge | -0.05 (-0.13, 0.03) | 0.22 | 0.22 |
| VidalBraloAge | -0.18 (-0.33, -0.02) | 0.03 | 0.04 |
| ZhangAge | -0.55 (-1.11, 0.00) | 0.05 | 0.06 |
| **Phenotypic age deviation** | | | |
| PhenoAge | -0.22 (-0.34, -0.11) | 0.001 | 0.002 |
| GrimAgeMort | -0.43 (-0.65, -0.21) | 0.0007 | 0.002 |
| GrimAge2Mort | -0.42 (-0.63, -0.22) | 0.0005 | 0.002 |
| DunedinPoAm | -0.14 (-0.31, 0.03) | 0.09 | 0.10 |
| **Other** | | | |
| HorvathTelo | -2.26 (-3.46, -1.05) | 0.001 | 0.002 |
| YangCell | -1.83 (-2.55, -1.11) | 0.00006 | 0.0008 |

Models adjusted for age, sex, race/ethnicity, education, smoking status, serum cotinine, and BMI

Chronological and phenotypic age deviation models are per one-year change in age deviation. Other models are per one-standard deviation change.

P-values adjusted by controlling for the False Discovery Rate (FDR)

**Supplemental Table 5.** Summary of sex-stratified survey-weighted linear regression model results evaluating the association between DNA methylation measures and Digit Symbol Substitution Test scores in NHANES, 1999-2002 (N = 1,463).

| DNA methylation measure | Overall (N = 1,463) | Male (N = 725) | Female (N = 738) | Interaction p (adj.) |
|---|---|---|---|---|
| **Chronological age deviation** | | | | |
| HorvathAge | -0.14 (-0.25, -0.03)* | -0.28 (-0.42, -0.13)* | -0.03 (-0.19, 0.13) | 0.12 |
| HannumAge | -0.32 (-0.45, -0.19)* | -0.54 (-0.71, -0.37)* | -0.15 (-0.37, 0.08) | 0.07 |
| SkinBloodAge | -0.25 (-0.47, -0.04)* | -0.56 (-0.74, -0.39)* | 0.05 (-0.26, 0.36) | 0.02 |
| LinAge | -0.08 (-0.16, 0.00) | -0.19 (-0.34, -0.05)* | 0.02 (-0.15, 0.20) | 0.19 |
| WeidnerAge | -0.05 (-0.13, 0.03) | 0.02 (-0.10, 0.15) | -0.10 (-0.21, 0.01) | 0.21 |
| VidalBraloAge | -0.18 (-0.34, -0.03)* | -0.22 (-0.45, 0.01) | -0.16 (-0.38, 0.06) | 0.75 |
| ZhangAge | -0.61 (-1.12, -0.09)* | -1.27 (-1.80, -0.75)* | 0.04 (-0.67, 0.74) | 0.06 |
| **Phenotypic age deviation** | | | | |
| PhenoAge | -0.22 (-0.33, -0.11)* | -0.38 (-0.52, -0.24)* | -0.10 (-0.30, 0.09) | 0.12 |
| GrimAgeMort | -0.42 (-0.64, -0.20)* | -0.56 (-0.81, -0.30)* | -0.30 (-0.58, -0.02) | 0.19 |
| GrimAge2Mort | -0.41 (-0.61, -0.21)* | -0.51 (-0.74, -0.28)* | -0.32 (-0.60, -0.05) | 0.26 |
| DunedinPoAm | -0.13 (-0.28, 0.03) | -0.24 (-0.46, -0.01) | -0.07 (-0.25, 0.11) | 0.22 |
| **Other** | | | | |
| HorvathTelo | -2.27 (-3.40, -1.14)* | -3.46 (-5.42, -1.50)* | -1.10 (-2.69, 0.49) | 0.18 |
| YangCell | -1.80 (-2.47, -1.12)* | -1.82 (-3.20, -0.45)* | -1.78 (-2.71, -0.85)* | 0.96 |

Models adjusted for age, sex, race/ethnicity, education, smoking status, serum cotinine ($log_2$ transformed and z-score standardized), and BMI, with an interaction term between the DNA methylation measure and sex. Chronological and phenotypic age deviation models are per one-year change in age deviation. Other models are per one-standard deviation change.

P-values adjusted by controlling for the False Discovery Rate (FDR).

* FDR-adjusted p < 0.05.

**Supplemental Table 6.** Summary of education-stratified survey-weighted linear regression model results evaluating the association between DNA methylation measures and Digit Symbol Substitution Test (DSST) scores in NHANES, 1999-2002 (N = 1,463).

| DNA methylation measure | Overall (N = 1,463) | Less than HS (N = 660) | HS or some college (N = 591) | College or above (N = 212) | Interaction p (adj.) |
|---|---|---|---|---|---|
| **Chronological age deviation** | | | | | |
| HorvathAge | -0.14 (-0.25, -0.03)* | -0.19 (-0.42, 0.05) | -0.09 (-0.27, 0.09) | -0.17 (-0.35, 0.02) | 0.97 |
| HannumAge | -0.32 (-0.45, -0.19)* | -0.33 (-0.65, -0.02) | -0.34 (-0.55, -0.13)* | -0.24 (-0.58, 0.11) | 0.97 |
| SkinBloodAge | -0.25 (-0.47, -0.04)* | -0.33 (-0.62, -0.04) | -0.19 (-0.59, 0.22) | -0.24 (-0.61, 0.13) | 0.97 |
| LinAge | -0.08 (-0.16, 0.00) | -0.02 (-0.20, 0.17) | -0.11 (-0.25, 0.03) | -0.12 (-0.34, 0.09) | 0.97 |
| WeidnerAge | -0.05 (-0.13, 0.03) | -0.02 (-0.17, 0.14) | -0.12 (-0.22, -0.02) | 0.04 (-0.14, 0.23) | 0.97 |
| VidalBraloAge | -0.18 (-0.34, -0.03)* | -0.16 (-0.52, 0.20) | -0.18 (-0.37, 0.01) | -0.24 (-0.71, 0.22) | 0.97 |
| ZhangAge | -0.61 (-1.12, -0.09)* | -0.90 (-1.53, -0.27) | -0.22 (-1.15, 0.72) | -0.68 (-1.68, 0.32) | 0.97 |
| **Phenotypic age deviation** | | | | | |
| PhenoAge | -0.22 (-0.33, -0.11)* | -0.26 (-0.51, -0.01) | -0.20 (-0.40, 0.00) | -0.21 (-0.52, 0.09) | 0.97 |
| GrimAgeMort | -0.42 (-0.64, -0.20)* | -0.28 (-0.71, 0.14) | -0.44 (-0.70, -0.17)* | -0.55 (-1.03, -0.07) | 0.97 |
| GrimAge2Mort | -0.41 (-0.61, -0.21)* | -0.33 (-0.74, 0.07) | -0.43 (-0.67, -0.20)* | -0.45 (-0.92, 0.02) | 0.97 |
| DunedinPoAm | -0.13 (-0.28, 0.03) | -0.13 (-0.44, 0.18) | -0.10 (-0.27, 0.07) | -0.20 (-0.60, 0.21) | 0.97 |
| **Other** | | | | | |
| HorvathTelo | -2.27 (-3.40, -1.14)* | -2.01 (-3.85, -0.17) | -2.47 (-4.19, -0.75) | -2.24 (-4.30, -0.17) | 0.97 |
| YangCell | -1.80 (-2.47, -1.12)* | -2.94 (-4.40, -1.48)* | -1.51 (-2.65, -0.37) | -0.60 (-2.85, 1.65) | 0.52 |

Models adjusted for age, sex, race/ethnicity, education, smoking status, serum cotinine ($\log_2$ transformed and z-score standardized), and BMI, with an interaction term between the DNA methylation measure and education.

Chronological and phenotypic age deviation models are per one-year change in age deviation. Other models are per one-standard deviation change.

P-values adjusted by controlling for the False Discovery Rate (FDR).

* FDR-adjusted p < 0.05.

**Supplemental Table 7.** Summary of race/ethnicity-stratified survey-weighted linear regression model results evaluating the association between DNA methylation measures and Digit Symbol Substitution Test (DSST) scores, NHANES 1999-2002 (N = 1,463).

| DNA methylation measure | Overall (N = 1,463) | Non-Hispanic White (N = 637) | Non-Hispanic Black (N = 278) | Mexican-American (N = 427) | Other Hispanic (N = 79) | Other Race (N = 42) | Interaction p (adj.) |
|---|---|---|---|---|---|---|---|
| **Chronological age deviation** | | | | | | | |
| HorvathAge | -0.14 (-0.25, -0.03)* | -0.15 (-0.27, -0.02) | 0.01 (-0.40, 0.43) | 0.23 (-0.56, 1.02) | -0.15 (-0.63, 0.33) | -0.09 (-0.57, 0.38) | 0.70 |
| HannumAge | -0.32 (-0.45, -0.19)* | -0.33 (-0.47, -0.19)* | 0.00 (-0.55, 0.56) | -0.08 (-1.19, 1.03) | -0.10 (-0.78, 0.59) | -0.45 (-1.33, 0.43) | 0.70 |
| SkinBloodAge | -0.25 (-0.47, -0.04)* | -0.27 (-0.49, -0.05) | 0.09 (-0.54, 0.72) | 0.13 (-1.23, 1.49) | -0.14 (-0.72, 0.45) | -0.20 (-0.75, 0.36) | 0.72 |
| LinAge | -0.08 (-0.16, 0.00) | -0.08 (-0.16, 0.00) | 0.10 (-0.40, 0.59) | 0.17 (-0.33, 0.68) | -0.10 (-0.61, 0.41) | 0.38 (-0.31, 1.06) | 0.83 |
| WeidnerAge | -0.05 (-0.13, 0.03) | -0.05 (-0.14, 0.03) | -0.13 (-0.57, 0.31) | -0.50 (-1.15, 0.15) | -0.24 (-0.64, 0.15) | 0.18 (-0.69, 1.05) | 0.70 |
| VidalBraloAge | -0.18 (-0.34, -0.03)* | -0.20 (-0.36, -0.03) | -0.09 (-0.59, 0.42) | 0.13 (-1.05, 1.31) | 0.21 (-0.30, 0.71) | -0.13 (-1.50, 1.24) | 0.70 |
| ZhangAge | -0.61 (-1.12, -0.09)* | -0.64 (-1.17, -0.11) | 0.65 (-0.87, 2.17) | 1.20 (-2.19, 4.59) | -0.23 (-1.49, 1.02) | -1.00 (-2.70, 0.71) | 0.70 |
| **Phenotypic age deviation** | | | | | | | |
| PhenoAge | -0.22 (-0.33, -0.11)* | -0.24 (-0.36, -0.12)* | -0.25 (-0.65, 0.15) | 0.45 (-0.30, 1.20) | -0.09 (-0.49, 0.30) | 0.58 (-0.43, 1.59) | 0.70 |
| GrimAgeMort | -0.42 (-0.64, -0.20)* | -0.43 (-0.66, -0.20)* | -0.50 (-1.34, 0.34) | -0.85 (-2.28, 0.57) | 0.10 (-0.48, 0.69) | 0.06 (-1.39, 1.50) | 0.70 |
| GrimAge2Mort | -0.41 (-0.61, -0.21)* | -0.43 (-0.64, -0.21)* | -0.50 (-1.18, 0.17) | -1.03 (-2.22, 0.16) | 0.00 (-0.54, 0.54) | 0.22 (-0.90, 1.33) | 0.70 |
| DunedinPoAm | -0.13 (-0.28, 0.03) | -0.13 (-0.30, 0.04) | -0.33 (-0.88, 0.22) | -0.34 (-1.17, 0.48) | 0.26 (-0.30, 0.83) | 0.05 (-1.24, 1.35) | 0.70 |
| **Other** | | | | | | | |
| HorvathTelo | -2.27 (-3.40, -1.14)* | -2.57 (-3.89, -1.26)* | -0.08 (-1.58, 1.41) | -0.15 (-2.65, 2.34) | 1.21 (-1.26, 3.68) | -4.21 (-8.24, -0.17) | 0.70 |
| YangCell | -1.80 (-2.47, -1.12)* | -2.01 (-2.76, -1.26)* | 0.07 (-1.83, 1.97) | -1.86 (-3.87, 0.15) | -0.90 (-3.32, 1.53) | -4.93 (-8.42, -1.45) | 0.70 |

Models adjusted for age, sex, race/ethnicity, education, smoking status, serum cotinine ($log_2$ transformed and z-score standardized), and BMI, with an interaction term between the DNA methylation measure and race/ethnicity.

Chronological and phenotypic age deviation models are per one-year change in age deviation. Other models are per one-standard deviation change.

P-values adjusted by controlling for the False Discovery Rate (FDR).
* FDR-adjusted p < 0.05.
